# Genomic perspectives on foodborne illness

**DOI:** 10.1101/2024.05.16.24307425

**Authors:** David J. Lipman, Joshua L. Cherry, Errol Strain, Richa Agarwala, Steven M. Musser

**Affiliations:** Center for Food Safety and Applied Nutrition, Office of Regulatory Science, College Park, MD, USA; National Center for Biotechnology Information, National Library of Medicine, National Institutes of Health, Bethesda, Maryland, USA; Division of International Epidemiology and Population Studies, Fogarty International Center, National Institutes of Health, Bethesda, Maryland, USA

## Abstract

Whole-genome sequencing of bacterial pathogens is used by public health agencies to link cases of food poisoning caused by the same source of contamination. The vast majority of these appear to be sporadic cases associated with small contamination episodes and do not trigger investigations. We analyzed clusters of sequenced clinical isolates of *Salmonella*, *Escherichia coli*, *Campylobacter*, and *Listeria* that differ by only a small number of mutations to provide a new understanding of the underlying contamination episodes. These analyses provide new evidence that the youngest age groups have greater susceptibility to infection from *Salmonella*, *Escherichia coli*, and *Campylobacter* than older age groups. This age bias is weaker for the common *Salmonella* serovar Enteritidis than Salmonella in general. Analysis of these clusters reveals significant regional variations in relative frequencies of *Salmonella* serovars across the United States. A large fraction of the contamination episodes causing sickness appear to have long duration. For example, 50% of the *Salmonella* cases are in clusters that persist for almost three years. For all four pathogen species, the majority of the cases were part of genetic clusters with illnesses in multiple states and likely to be caused by contaminated commercially distributed foods. The vast majority of *Salmonella* cases among infants < 6 months of age appear to be caused by cross-contamination from foods consumed by older age groups or by environmental bacteria rather than infant formula contaminated at production sites.

## Introduction

Since 2013, the CDC, FDA, and USDA have been using whole genome sequencing (WGS) of pathogens to detect foodborne outbreaks and to trace the source of contamination (Jackson et al. 2016). Because isolation of the pathogen and subsequent whole genome sequencing (WGS) has become routine for reportable illnesses, we now have comprehensive sequence databases of *Salmonella*, *Escherichia coli*, *Campylobacter*, and *Listeria* genomes. While the use of WGS has allowed CDC and FDA to detect foodborne outbreaks earlier and to improve the success rate for identification of the contamination source(B. Brown et al. 2021), the associated sequence databases also provide a new way to study sporadic cases of foodborne illnesses that are far more prevalent than outbreak cases <u>(Ebel et al. 2016)</u>.

If the genome sequences of a set of clinical isolates differ by only a small number of mutations (e.g. by ≤ 4 single nucleotide polymorphisms, SNPs, for the entire genome sequence), they are very likely to share the same source of contamination <u>(Brown et al. 2019; Jagadeesan et al. 2019; Ronholm et al. 2016;</u> <u>Stevens et al. 2022; Pightling et al. 2018)</u>. This is the central basis for using WGS in food safety. The contamination can occur at any stage of the food production, distribution, or preparation process, including within the consumer’s household. For the purposes of this analysis, we will define a “contamination episode” as the set of contamination events caused by a single source. The term “episode” is being used because the contamination events from this source may occur over a period of time.

The pathogen genomes in CDC’s PulseNet database (Tolar et al. 2019) isolated from clinical cases of foodborne illness represent a large subset of the servings of food impacted by contamination episodes (E. Brown et al. 2019). These are the cases where:

● An individual consumed a serving of food with a high enough level of contamination to cause sufficient illness to seek medical care;

○ Note that the level of contamination needed to cause illness can vary related to patient and pathogen factors;
● The laboratory tests had to be sufficiently sensitive to pick up the reportable pathogen;
● The patient sample had to be submitted to the state public health lab for sequencing.

A number of estimates have been made for the actual number of cases of foodborne illness from the major pathogens, i.e., the multiplier for the reported cases. As an example, CDC estimates that the actual number of cases due to *Salmonella* is approximately 29 times the number of reported cases (Scallan et al. 2011; Thomas et al. 2015). In larger contamination episodes, many food servings may be contaminated at levels sufficient to cause illness in most consumers but other affected servings may have relatively small levels that have a low probability of causing symptoms in healthy adults. However, in more susceptible individuals (e.g. young children and the older adults), these levels may have a higher probability of causing disease. In countries with modern food production systems and active safety programs, such as the United States, one might expect most contamination episodes to be small - impacting only a small number of servings and with infectious doses that have only a low probability of causing illness in most individuals. These smaller contamination episodes are called sporadic food poisoning and are known to be responsible for the vast majority of foodborne illness (Ebel et al. 2016).

Sporadic food poisoning, because it does not trigger an epidemiological investigation, has largely been studied by case-control studies. Because the genome sequences now allow us to identify clusters of cases associated with a contamination episode, we can analyze these contamination episodes more directly. In other words, the whole genome sequencing data helps us to connect the genetic clusters of clinical cases to the underlying contamination episodes. In addition, we can compare trends among different species and serovars of foodborne pathogens as well as in different regions across the United States and make inferences regarding the contamination episodes associated with clinical cases of food poisoning. Note that the approach described here does not fully account for contamination episodes that are polyclonal: each of the strains in polyclonal contamination episodes will appear as a single cluster (Sarno et al. 2021; Gerner-Smidt et al. 2019).

By examining the composition of these clusters, we can obtain estimates of the fraction of contamination episodes occurring upstream of the distribution from a central source of production because clinical cases from the same cluster are found in multiple states. Likewise, by using the isolation dates of the clinical cases in a cluster, we can observe the persistence of the contamination episode. Finally, by using the ages of the members of a contamination cluster, we can get important clues as to whether the serving of food was contaminated prior to entering the household or whether there was cross-contamination of the serving from other foods in the household.

## Results

The Methods section describes how the clusters of clinical cases of food poisoning are generated. Briefly, we use a threshold genetic distance for pairs of genomes that is low enough to infer a high likelihood that the associated pathogen isolates are derived from the same source of contamination, e.g., 4 SNPs out of ∼2-4 million aligned bases. We continue to add cases to the cluster as long as they are within this threshold distance from at least one case already in the cluster, i.e., we perform single linkage clustering.

The results presented here are based on a 4 SNP threshold but using smaller or larger thresholds did not qualitatively change the results (see Methods).

Figure 1 is a cumulative frequency plot of the percent of all cases in clusters of size 1 on up to size 30 for *Salmonella*, *Escherichia coli*, *Campylobacter*, and *Listeria*. For *Salmonella*, we separated out Enteritidis isolates because Enteritidis appears quite different from all the other major serovars and the other pathogen species. Other than Enteritidis, the four pathogen species have over 50% of all cases in singleton clusters, i.e., these cases are more than 4 SNPs away from all other cases, and over 75% of the cases are in clusters of size ten or less. For Enteritidis, less than 20% of the cases are in singleton clusters (see Figure 1) and 50% of the cases are in clusters larger than size 50 (data not shown).

**Figure 1.**
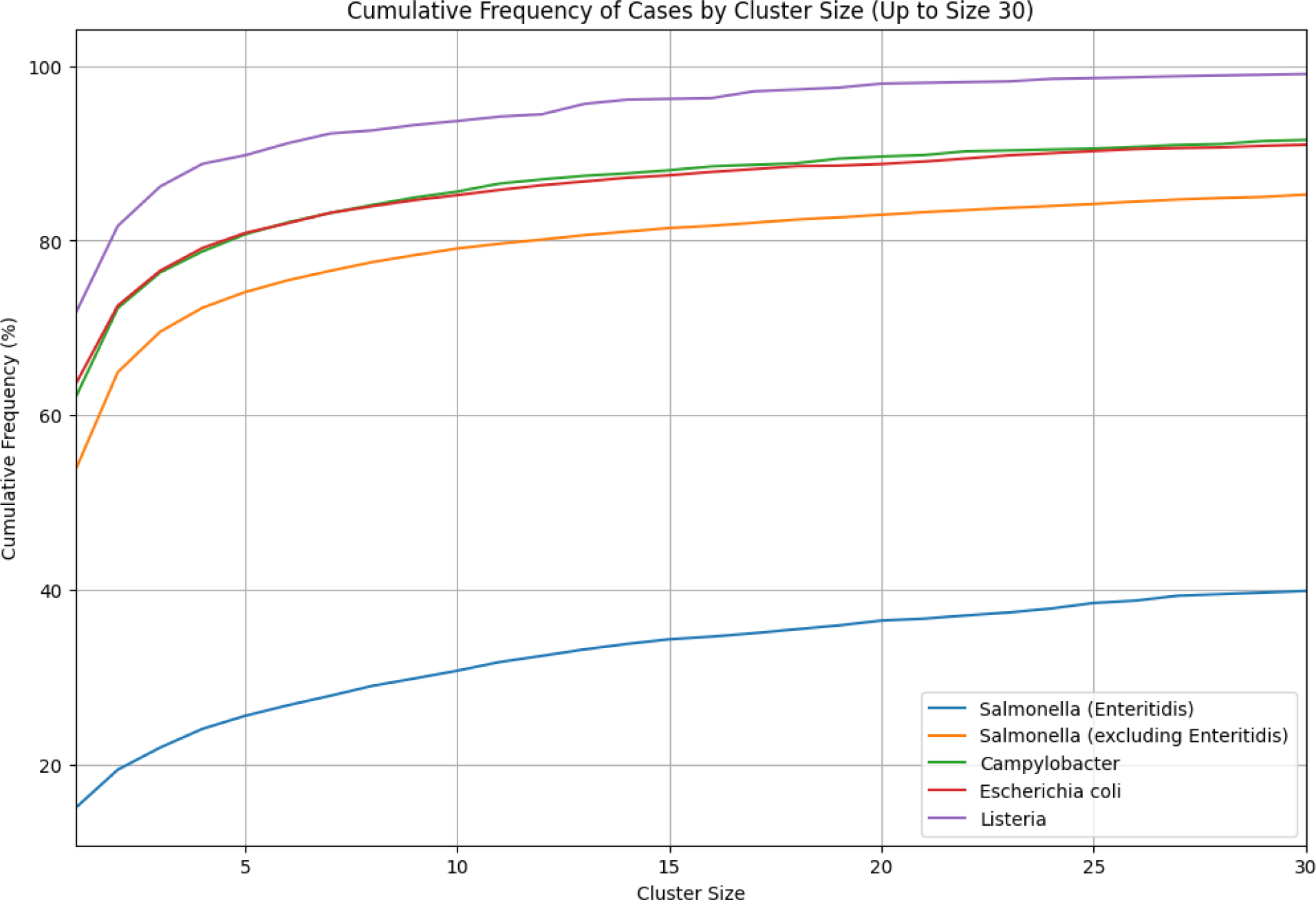
Cumulative frequency of cases by cluster size Cumulative frequency plot showing the percent of all cases in clusters of size 1 up to size 30 for *Salmonella*, *Escherichia coli*, *Campylobacter*, and *Listeria*. Enteritidis isolates are separated from other *Salmonella* serovars due to their distinct clustering behavior.

We will examine some of the properties of clusters of different sizes to determine whether there is a clear signal associated with the computed clusters. Figure 2 shows the age distribution of the cases for all four pathogens in small clusters (size <=4) compared with larger clusters (size >20). The case counts have been normalized for the age population structure (“United States Population by Age and Sex” n.d.).

**Figure 2.**
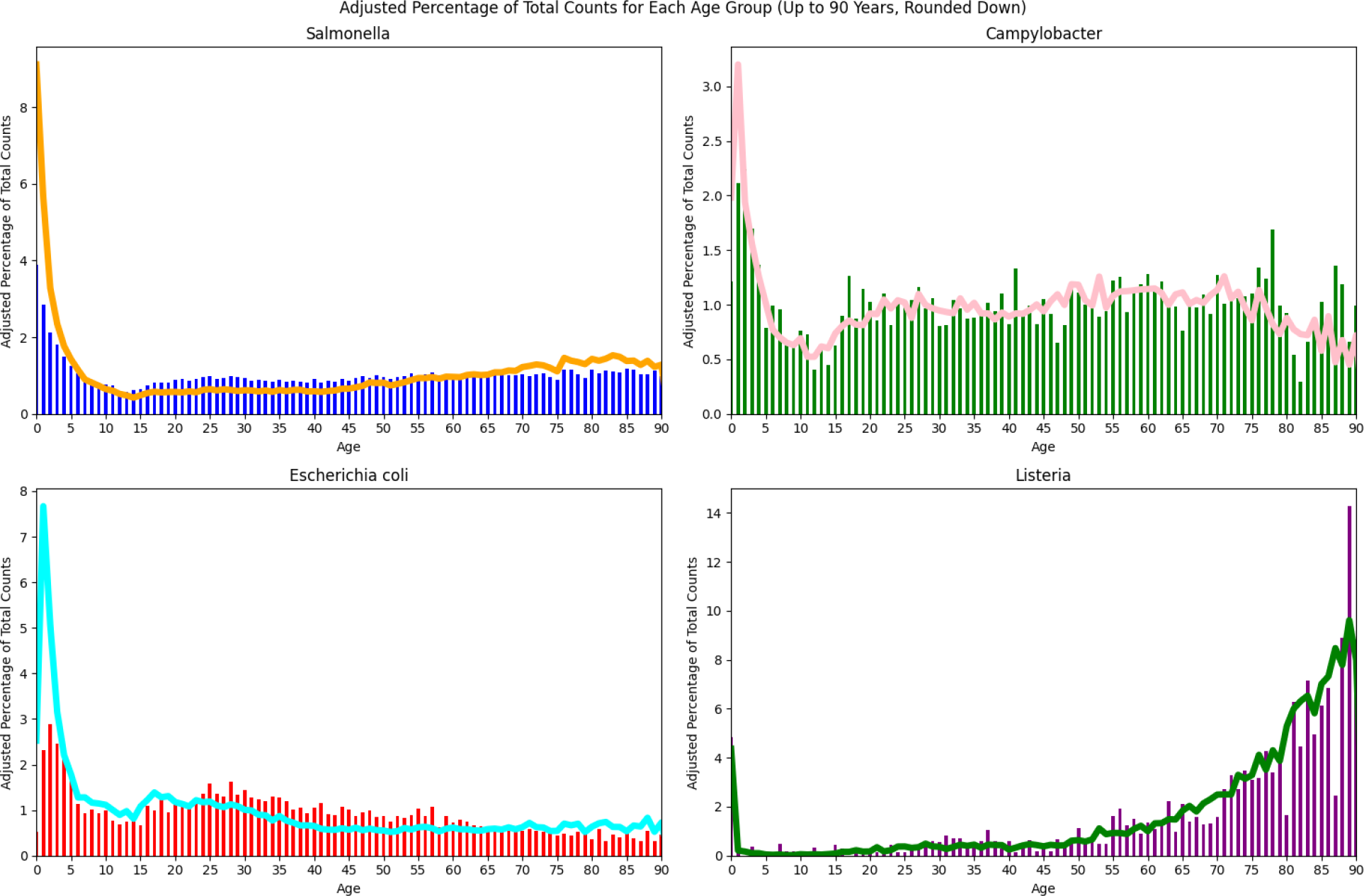
Age distribution of cases in small (<= 4) versus large clusters (>20) Age distribution of cases in small (cluster size ≤ 4, line) versus large clusters (cluster size > 20, vertical bars) across all four pathogens. The case counts have been normalized based on the US population for different age groups.

Particularly for the smaller clusters, *Escherichia coli* and *Campylobacter* show an increasing number of cases from approximately age 10 down to age 1 before a drop for infants under 1 year old. *Salmonella* is similar, though case counts appear to be increasing through age 0. However, nonmonotonicity becomes apparent when ages less than one year are binned more finely (Supp. Figure 1). Thus, the incidence of *Salmonella* also decreases as age approaches zero, but the downturn occurs at less than one year of age and is not apparent in Figure 2. The age distributions for *Salmonella* in Figure 2 are consistent with those reported in published surveillance studies though the latter do not break down into e.g. large and small cluster sizes cf (Boore et al. 2015). For *Salmonella*, *Escherichia coli*, and *Campylobacter*, the increased incidence in the youngest age groups is especially pronounced in the small cluster sizes, i.e., there is a substantially higher fraction of young individuals in the smallest clusters.

For *Salmonella* there is a subtle increase in older adult cases while for *Escherichia coli*, there is a small increase in the 18-30 year age interval followed by a decrease to a plateau from age 40 onwards. For *Salmonella*, *Escherichia coli*, and *Campylobacter* we see a trough in case counts between the infant/young child peak and the teenage years. *Listeria* looks quite different with case counts increasing with age along with a narrow peak associated with newborn Listeriosis (Charlier, Disson, and Lecuit 2020).

Among the major *Salmonella* serovars, Enteritidis stands out in having a far higher percentage of its cases in large clusters (Figure 1) and it accounts for 40% of the *Salmonella* cases in clusters larger than size 20 (data not shown). Furthermore, as seen in Figure 3 as compared with Figure 2a, the age distribution of Enteritidis is less skewed towards younger age groups and this difference is only somewhat more pronounced in small clusters.

**Figure 3.**
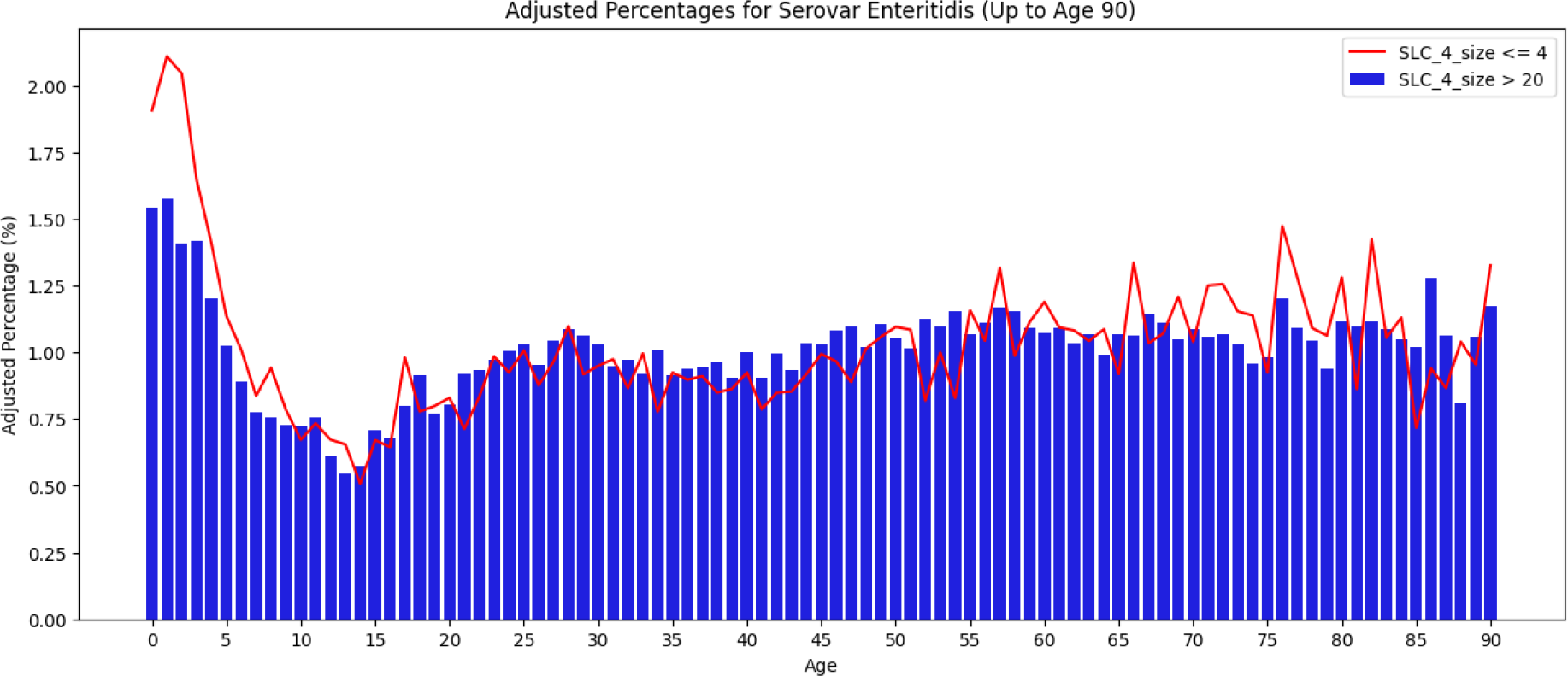
Age distribution of cases in small (<= 4) versus large clusters (>20) for *Salmonella* Enteritidis Age distribution of *Salmonella* Enteritidis cases in small (cluster size ≤ 4, line) versus large clusters (cluster size > 20, vertical bars). The case counts have been normalized based on the US population for different age groups.

Another property associated with cluster size is the diversity in terms of, e.g., serovars observed with the different cluster sizes. There are a variety of diversity measures used in ecology and in microbiology (Kim et al. 2017) with different emphases on, e.g., species richness vs the evenness of different species, or in our case, e.g., serovars. The boxplots in Figure 4a show the range of values for the Simpson Diversity Index (1-D) of cluster sizes 1 to 10 for the serovars of *Salmonella* (excluding Enteritidis). The resampling analysis used a fixed sample size of 1000 cases and 200 random samples for each cluster size. Similar results were obtained with the Shannon Diversity Index as well as by simply counting the number of unique serovars (data not shown). Thus there are a large number of relatively rare serovars and these are more commonly seen in the smaller clusters.

**Figure 4.**
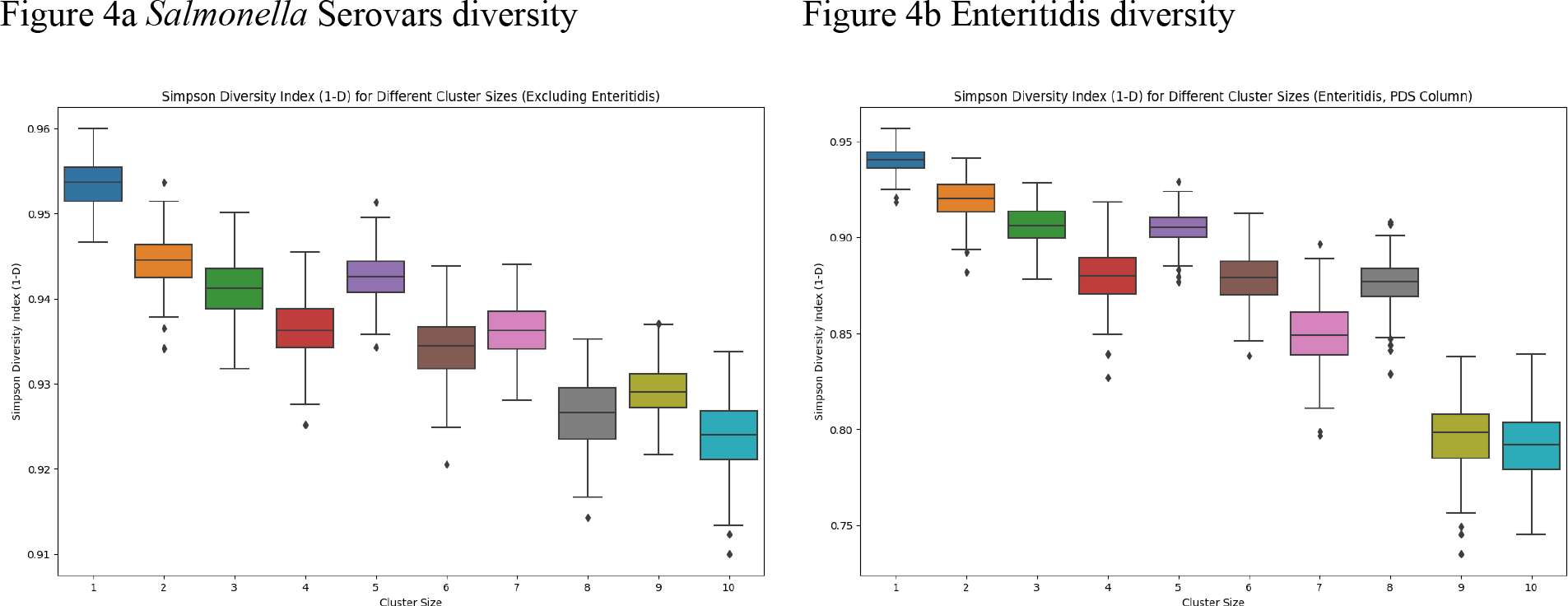
Diversity by size of cluster a) Resampling analysis of the diversity (Simpson diversity index 1-D) of *Salmonella* serovars for cluster sizes 1: 10. The resampling analysis used a fixed sample size of 1000 cases and 200 random samples for each cluster size. b) Resampling analysis of the diversity of Enteritidis within different NCBI PDS clusters for cluster sizes of 1:10. Sample size = 300, 200 random samples for each cluster size.

As an alternative to serovar, another measure of relatedness would be genetic distance among isolates. The NCBI Pathogen Detection SNP (PDS) clusters include all genomes that are within 50 SNPs of each other (see NCBI pipeline summary https://ftp.ncbi.nlm.nih.gov/pathogen/Methods.txt) and thus group closely related strains but also can include isolates from many unrelated contamination episodes. Figure 4b shows a resampling analysis of the Simpson Diversity Index by cluster size for Enteritidis analogous to Figure 4a but using the PDS clusters instead of serovar (sample size = 300, 200 random samples for each cluster size). Analyses using the PDS clusters for *Escherichia coli* and for *Campylobacter* yielded similar results as did using the Shannon Diversity Index or by simply counting unique NCBI clusters (data not shown). Thus there is a consistent trend towards greater diversity in the smaller cluster sizes, further evidence that these clusters are associated with meaningful differences in epidemiologically relevant properties.

Given the evidence from Figure 4 that there is more serovar diversity in the smaller clusters, a possible explanation for the age distribution results seen in Figure 2 might be that the younger age groups are more susceptible to and/or more frequently exposed to the serovars seen primarily in the smaller clusters.

Figure 5 compares the ratio of the number of cases in small clusters (<= 4) to the number in large clusters (> 20) by age group for common serovars (> 1.5% of cases), rare serovars (< 1.5% of cases), and for Enteritidis. The age ranges were chosen to include approximately the same numbers of cases and the ratio has been normalized because Enteritidis, unlike the other *Salmonella* serovars, has more cases in the large clusters than the small clusters. We see the same pattern for all three categories of serovars, i.e. there is a higher fraction of younger cases and of older cases in the smaller clusters. This same pattern holds for each of the common serovars (those accounting for > 1.5% of the cases) individually (data not shown). Thus, the higher frequencies of younger cases in small clusters seen in Figure 2a (and to a lesser extent, older cases) do not seem to be primarily due to the mix of serovars.

**Figure 5.**
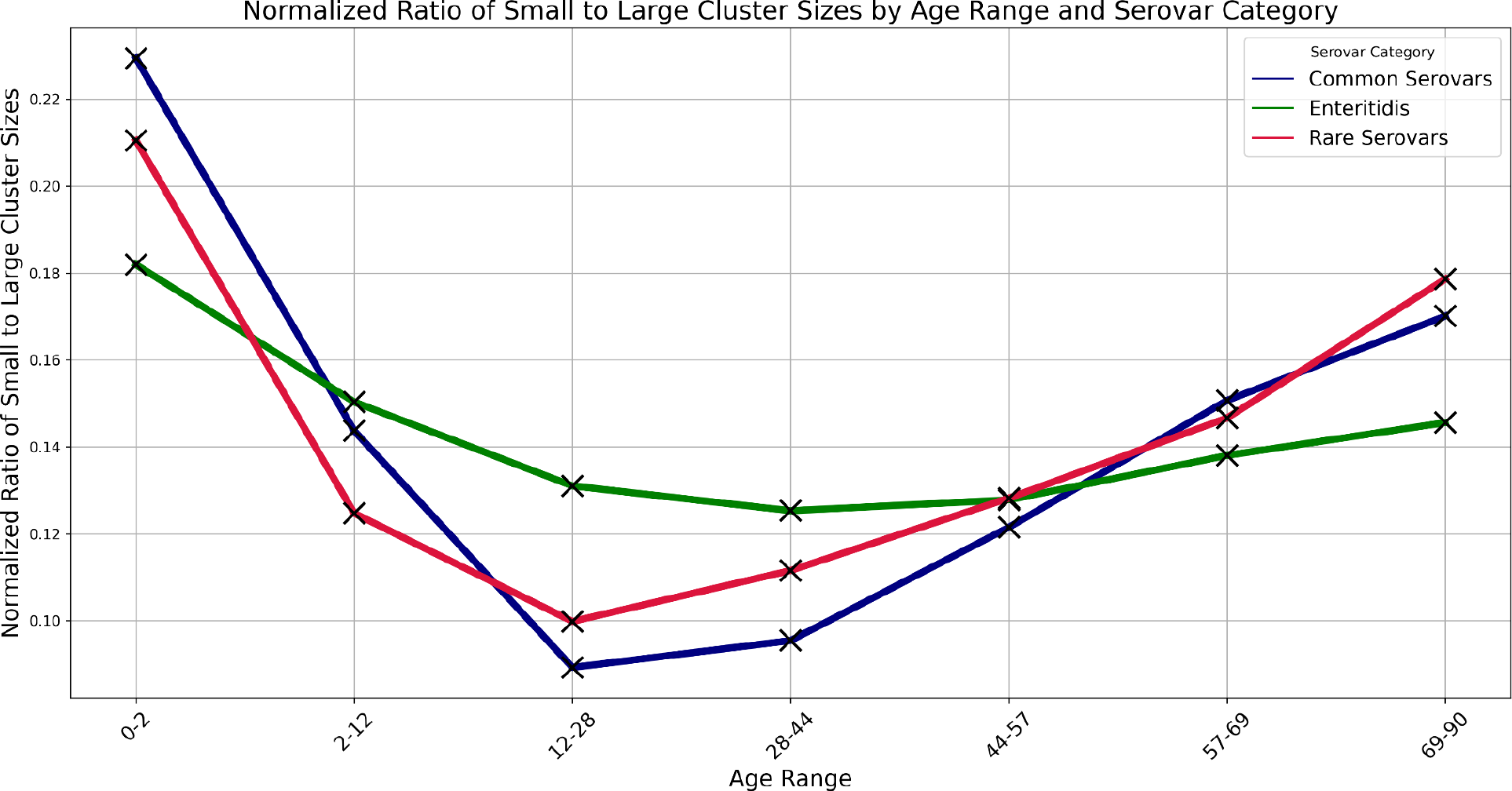
Ratio of small to large cluster sizes by age range and serovar category Ratio of the number of cases in small clusters (≤ 4) to the number in large clusters (> 20) by age range and serovar category. Common serovars were those that account for > 1.5% of the cases, excepting Enteritidis which is plotted separately. Rare serovars were those accounting for < 1.5% of the cases. The ratios for each serovar were normalized by summing across age ranges and dividing the ratio for a given age range by this sum.

Supplementary Figure 2 compares the singleton cluster case counts for each *Salmonella* serovar in infants (under one year old) to that in those at least ten years old. Figure 6 shows the same information for serovars with fewer than 2000 cases in the older age category. The gray line corresponds to equality of fraction of cases in the two age groups (not to equal case rates, as case rates are higher on average in infants). There is considerable variation in the ratio of infant to older cases among serovars. A small fraction of rare (in the older category) serovars are especially biased toward infants. Subspecies IV (red points), which is mostly adapted to cold-blooded vertebrates, is overrepresented among these. Overall, subspecies IV constitutes only 0.9% of the older cases but 6.8% of infant cases. Subspecies I, the main source of human infections, is also represented, but most prominently by serotypes that are associated with reptiles or other environmental sources, such as Rubislaw, Gaminara, and Poona.

**Figure 6.**
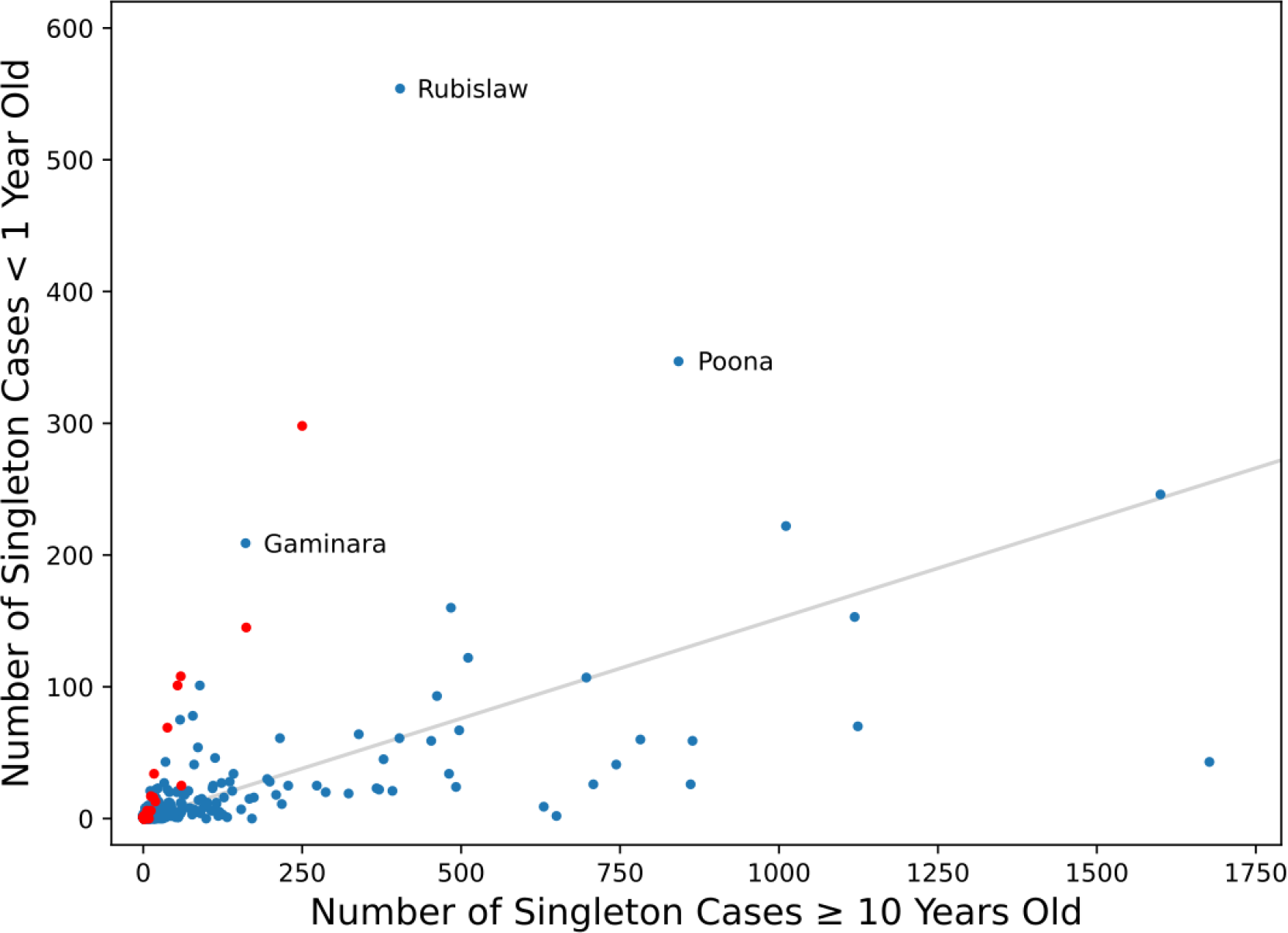
Comparison of *Salmonella* serovar occurrence in singleton clusters in infants and those at least ten years old. Types belonging to subspecies IV are shown in red. The gray line corresponds to equal fractions in the two age groups. Only serovars with fewer than 2000 cases in the older age group are shown.

Considering the geographical distribution of *Salmonella* cases (Figure 7a), we see that Enteritidis comprises a far higher percentage of the *Salmonella* cases in the Northeastern United States, stretching west to North Dakota while comprising a much lower percent of *Salmonella* cases in the Southeastern United States and Hawaii. This is consistent with the published CDC surveillance report (“2016-*Salmonella*-Report-508.pdf,” n.d.). While serovars Javiana and Newport are somewhat higher in the Southeast, the most dramatic difference is the much lower fraction of Enteritidis in the Southeastern United States and Hawaii. This is consistent with the FoodNet data (“FoodNet” 2023).

**Figure 7.**
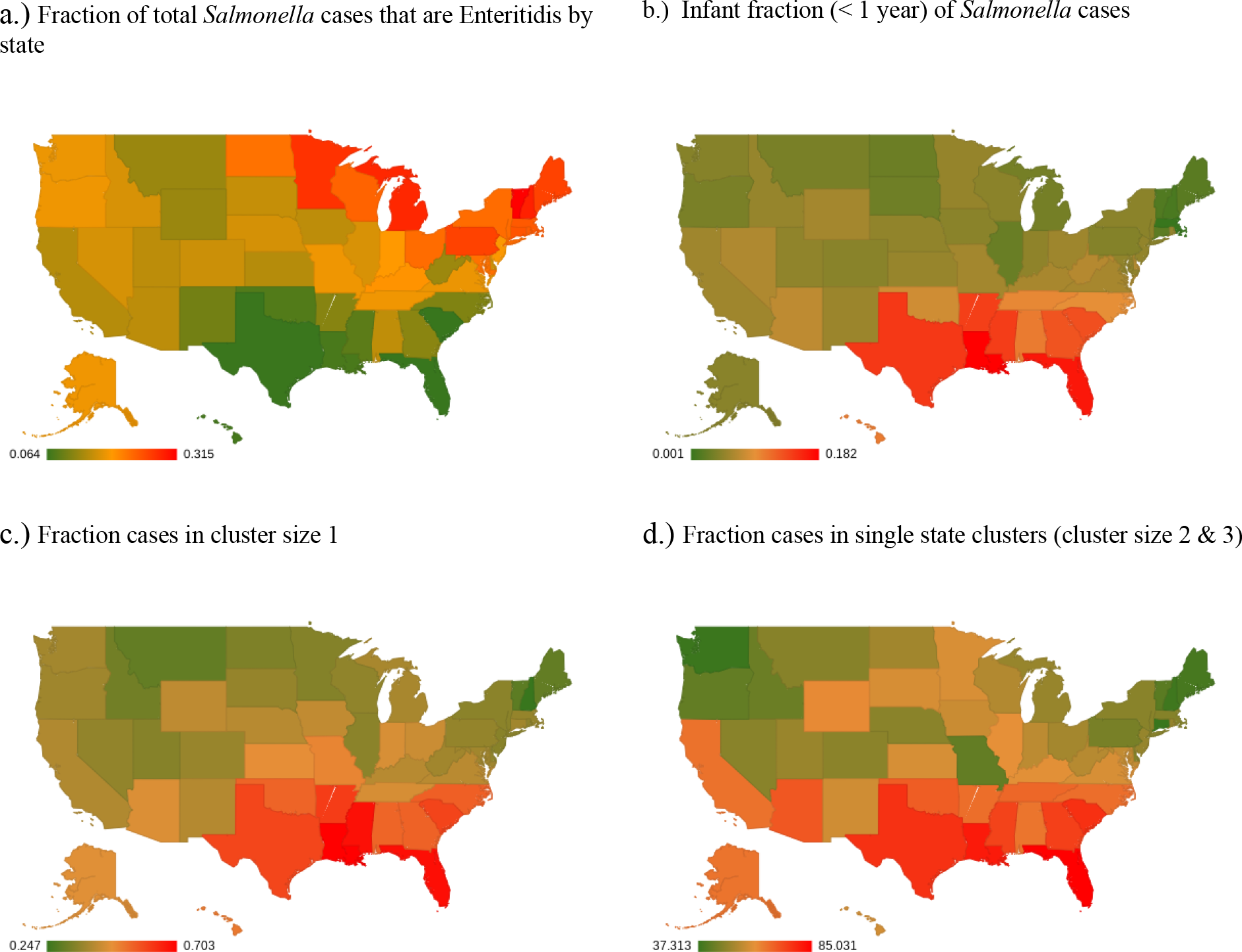
Geographical distribution of *Salmonella* cases Geographical distribution of *Salmonella* cases: a) the fraction of Enteritidis cases in each state, b) the fraction of infant cases (< 1 year), c) the fraction in clusters of size 1, d) the fraction of cases in single state clusters (cluster size 2 & 3).

[Centers for Disease Control and Prevention (CDC). FoodNet Fast: Pathogen Surveillance Tool. Atlanta, Georgia: U.S. Department of Health and Human Services. Available from URL: http://wwwn.cdc.gov/foodnetfast.] A higher fraction of infant cases (Figure 7b), cases in clusters of size 1 (Figure 7c), and single-state clusters (i.e., all members of a cluster are from the same state) (Figure 7d) are also seen in Southeastern states and Hawaii.

If this geographic distribution were due to the warmer and more humid climate of the Southeastern US states and Hawaii, we might expect a seasonal pattern of age incidence, with a higher fraction of infant cases in the warmer months. Figure 8 shows that although overall *Salmonella* incidence is seasonal, as observed previously (“FoodNet” 2023), the fraction of infant cases is fairly constant throughout the year. Thus, the geographic pattern does not seem to be a simple consequence of temperature.

**Figure 8.**
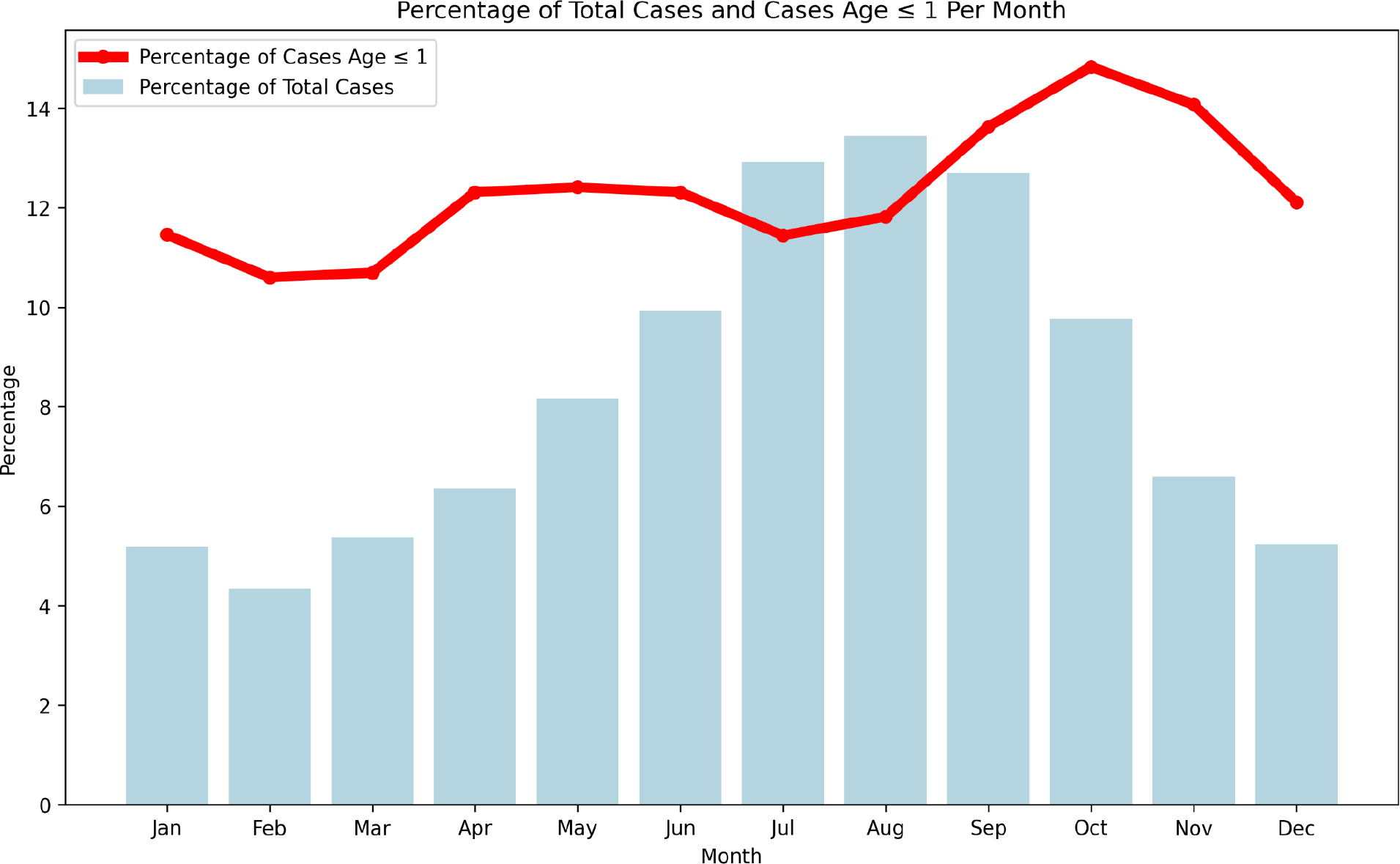
S*a*lmonella monthly cases vs monthly infant fraction (< 1 years old) Comparison of monthly *Salmonella* cases (vertical bars) versus the monthly fraction of infant cases (< 1 year old, line).

Because we know the dates of collection of the clinical isolates, we can examine the persistence of the clusters greater than size 1, i.e., the number of days between the first isolate collected and the most recent isolate in the cluster. Figure 9 shows the cumulative distribution of cluster persistence time among cases. Median persistence times are highest for *Salmonella* and lowest for *Escherichia coli*. Note that we are underestimating the persistence of clusters beginning or ending outside of the dates of our sample collection. An even greater number of cases are likely to be missed because, as mentioned in the Introduction, they have escaped detection by PulseNet surveillance.

**Figure 9.**
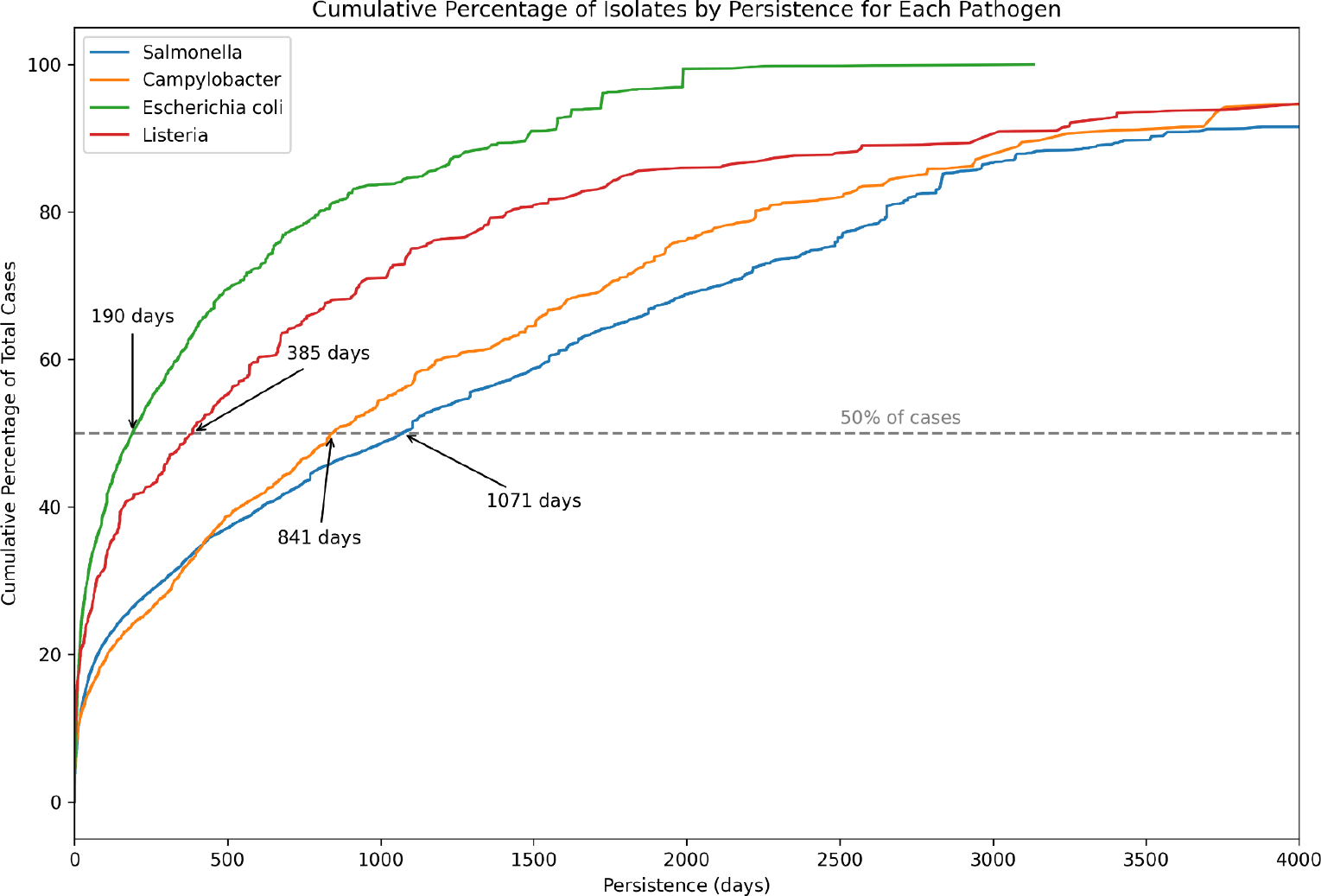
Persistence of clusters Cumulative Distribution of Cluster Persistence: The cumulative percentage of cases within non-singleton clusters, showing the range of cluster persistence measured by the number of days from the first to the last isolate collected within each cluster. The graph highlights the distribution of persistence times across all observed clusters, with a dashed horizontal line indicating the median persistence level, where 50% of cases are found in clusters with a duration exceeding this value.

We can also examine non-singleton clusters to see whether multiple US states are represented within them (Table 1). While some contamination episodes are likely from a source in the local environment, a majority of the contamination episodes appear to be geographically dispersed. That is, the contaminated servings of food are likely to have been distributed from a central point source (e.g., a food production or packaging facility) because cases from multiple states are members of the same cluster. Geographical dispersion is likely to be underestimated for the same reasons noted above for cluster persistence.

**Table 1.**
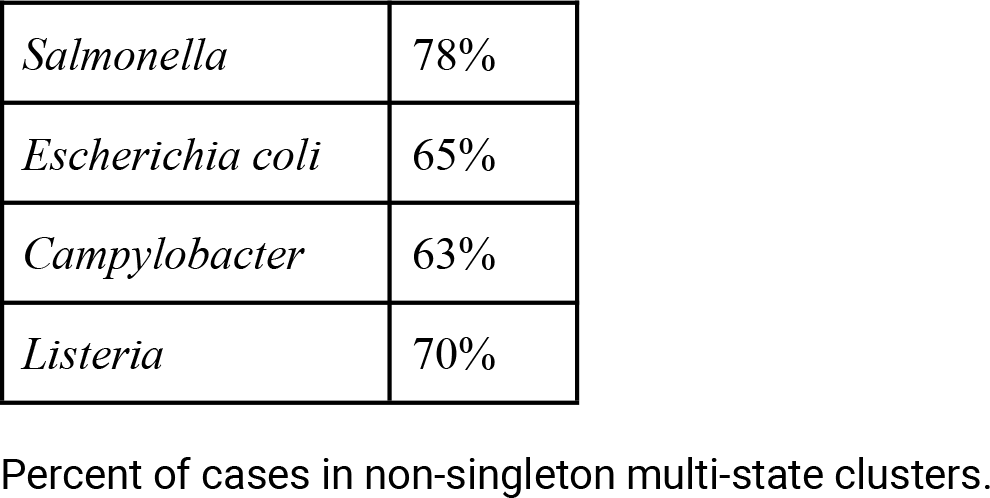
Percent of cases in multi-state clusters.

|  |  |
| --- | --- |
| *Salmonella* | 78% |
| *Escherichia coli* | 65% |
| *Campylobacter* | 63% |
| *Listeria* | 70% |

Percent of cases in non-singleton multi-state clusters.

Larger clusters are more likely to be multi-state for the trivial reason that, with more cases, there is a greater chance to include a case from a different state. If we examine all pairs of cases within clusters of a given size, we can see whether larger contamination episodes are inherently more likely to be distributed out from a central site rather than, e.g., acquired from some local environmental source (red line in Figure 10). For comparison, the blue line in Figure 10 shows the fraction of clusters with cases from at least two different states.

**Figure 10.**
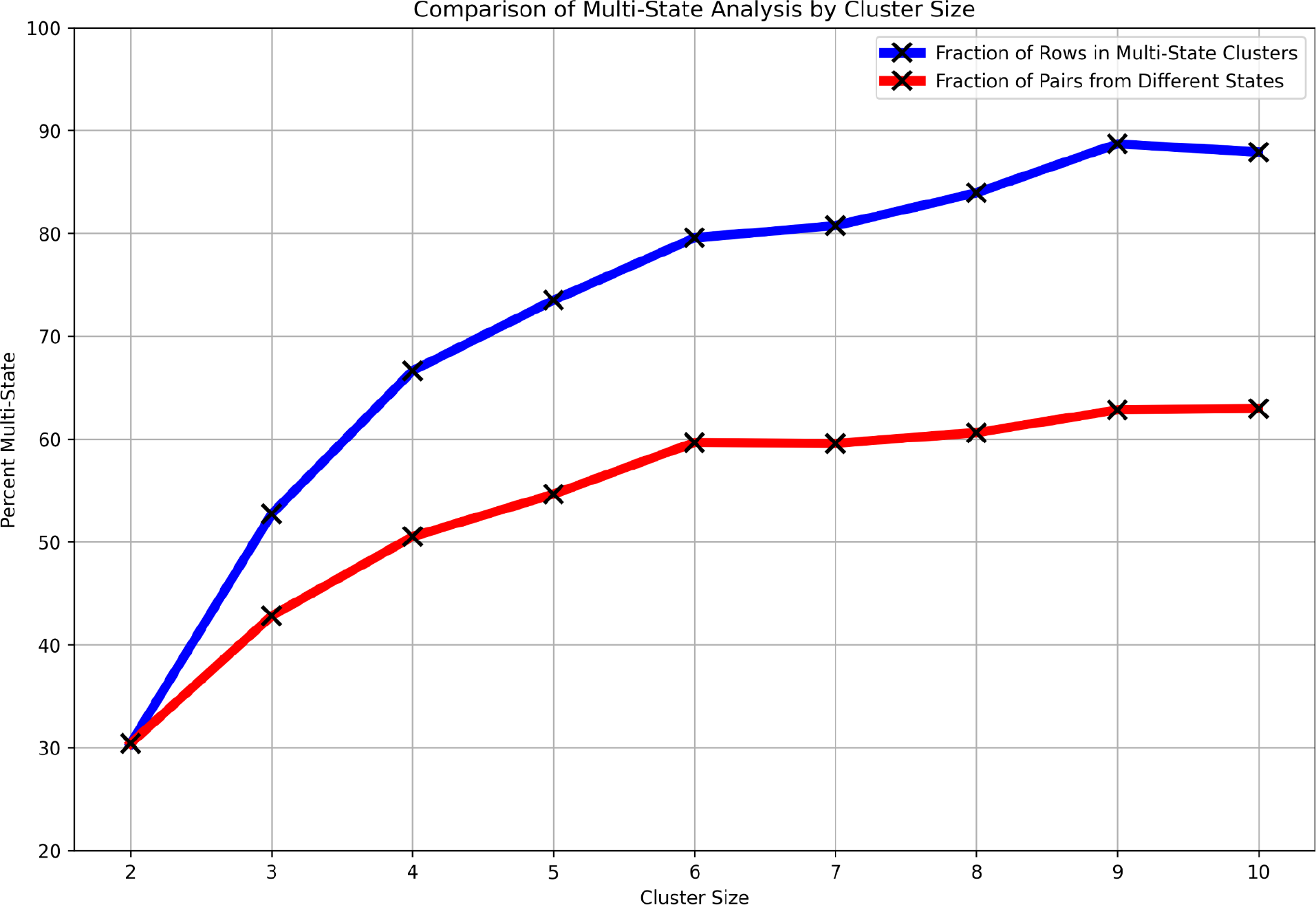
Fraction of multi-state *Salmonella* clusters vs cluster size Multi-state analysis by cluster size: The blue line shows the percent of cases in multi-state clusters for cluster sizes 2:10. The red line shows the fraction of pairs of cases in each cluster size that are from different states.

The red line shows that the contamination episodes underlying the larger clusters are inherently more likely to be distributed from a central site. While larger clusters are more likely to be geographically dispersed than smaller clusters, for *Salmonella*, over 30% of the clusters of size 2 are multi-state and by cluster size 3, over 50% are multi-state (Figure 10, blue line). Similar results were obtained for *Listeria*, *Campylobacter*, and *Escherichia coli* (data not shown).

Figure 11 compares the persistence of single state versus multi-state clusters for all four pathogens. We restricted the analysis to clusters of size 2 or 3 since the fraction of single state clusters is substantially lower with increasing cluster size (see Figure 10). As noted above, while the contamination for some of the single state clusters may be distributed out from e.g. a central site, a substantial fraction are likely to be local in nature. Most of the multi-state clusters, however, are likely to have been distributed from central sources. The degree of shift to higher persistence of the multi-state clusters compared to single-state appears roughly similar for all four pathogens.

**Figure 11.**
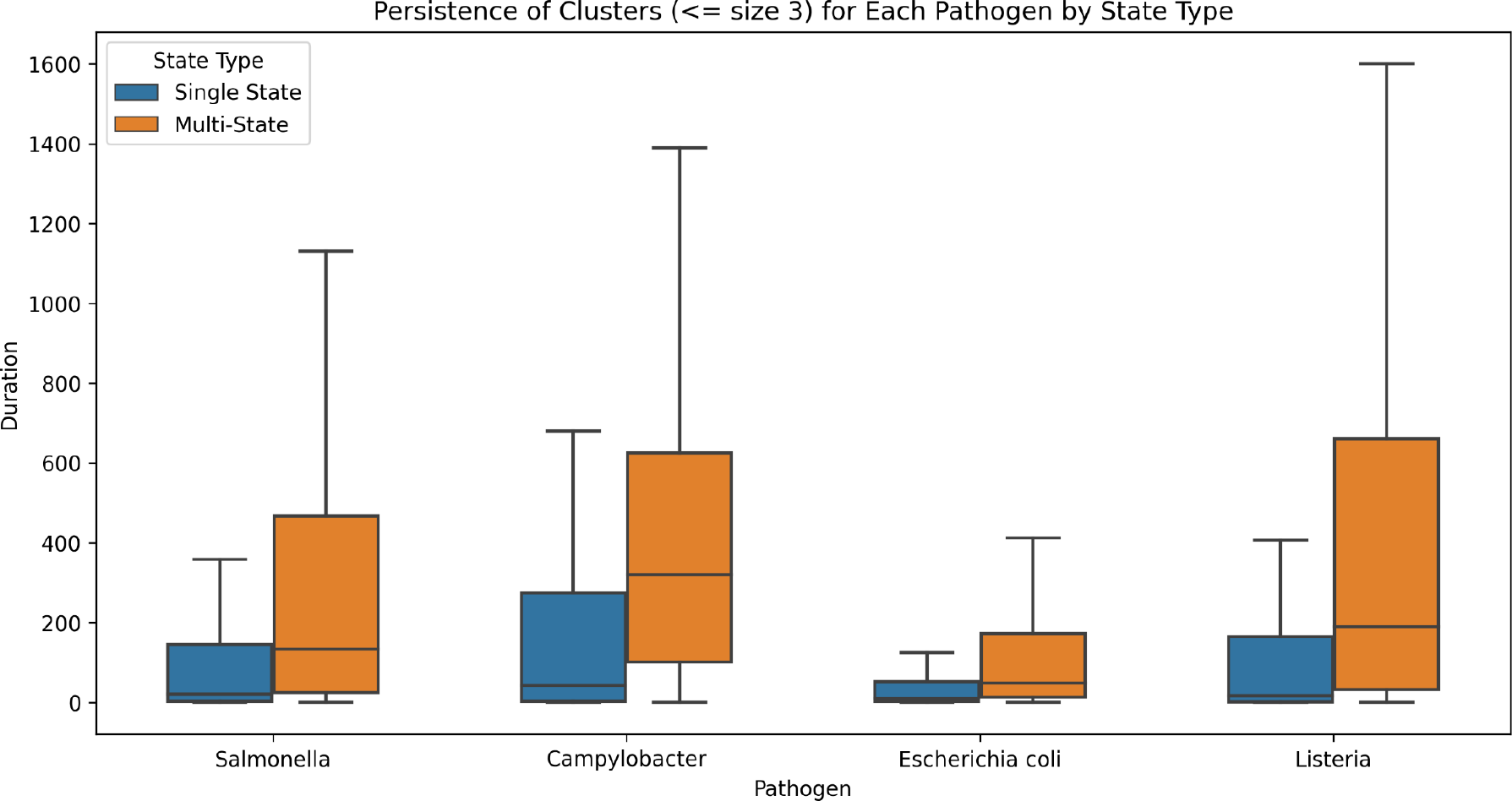
Comparing persistence of single state versus multi-state clusters Comparison of the persistence of single-state versus multi-state clusters of size 2 and size 3 for all four pathogens.

We can also examine how the geographical dispersion of cases within a cluster varies among age groups. The blue bars in Figure 12 show the percent of single-state clusters (size ≥ 2) for different age groups.

**Figure 12.**
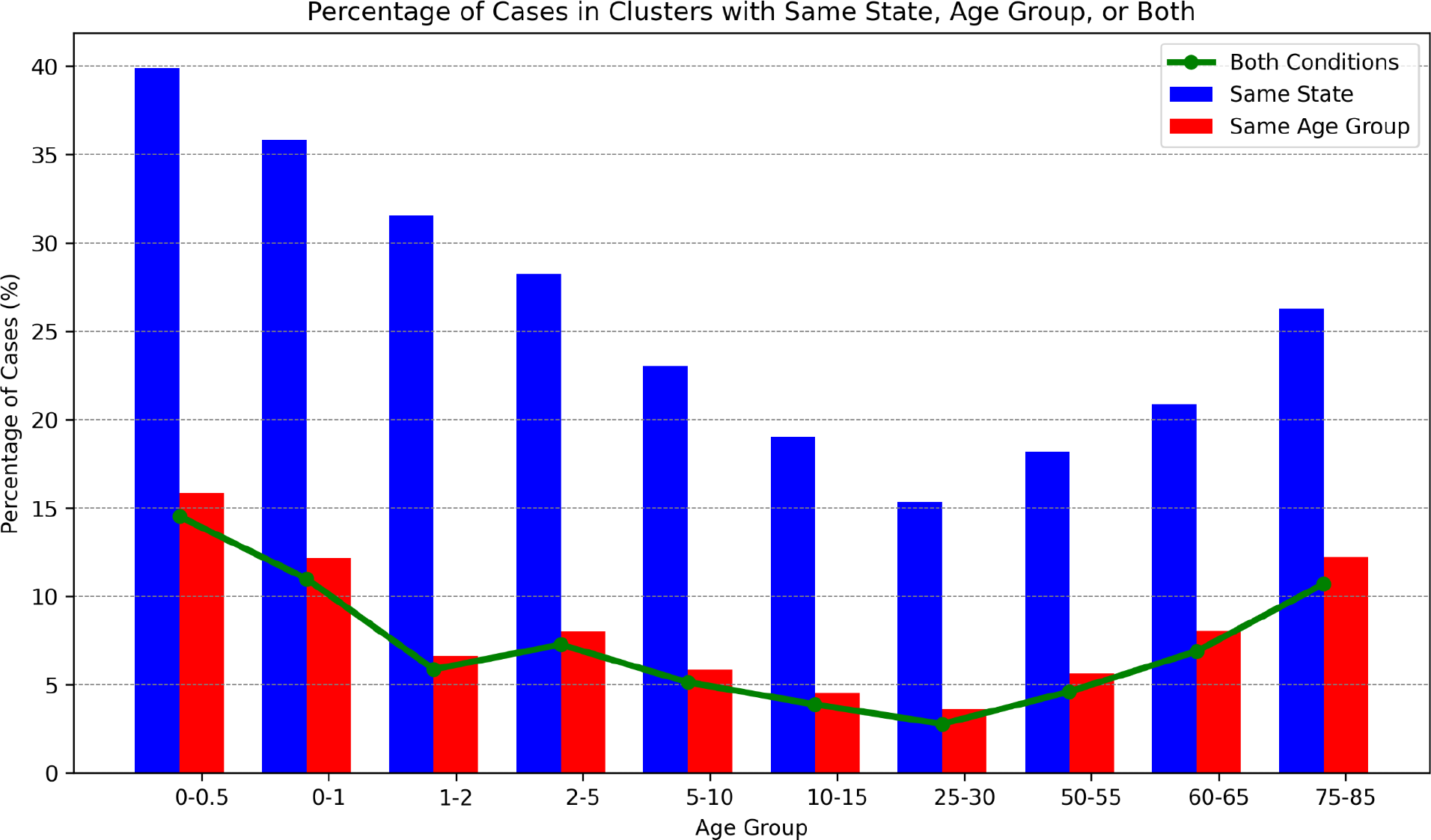
Case percentages in single-state clusters, single age-group clusters, and single-state + single age-group clusters Case percentages in single-state clusters (blue bars), single age-group clusters (red bars), and clusters that are both single-state *and* single-age-group (green line).

The youngest and oldest age groups have the highest percentage of cases in single-state clusters, which is consistent with their skew towards smaller clusters (Figures 2 and 5) and the relationship between cluster size and geographical dispersion (Figure 10). The red bars show the percent of cases in clusters where all cases in the cluster are within the same age range and this correlates very well with the geographical dispersion. The green line shows the percent of cases in clusters where all cases are in the same state *and* same age range - virtually all same-age clusters are also single-state. Thus, although overall only 22% of *Salmonella* isolates are in single-state (Table 1) clusters, *contamination episodes resulting in same-age clusters are virtually all single-state.* This would be consistent with these contamination episodes occurring in settings such as elder care facilities, daycare facilities, schools, and direct environmental exposure.

The diet of the age group less than 6 months old is distinct from all other age groups in that it consists almost entirely of breast milk and/or infant formula (Chiang et al. 2023). Furthermore, infant formula is distributed nationally from a small number of production facilities. If infant formula were a significant source of *Salmonella* contamination then one might expect to see a substantial fraction of multi-state clusters solely composed of infants rather than the very small fraction seen in Figure 12 (i.e., the distance between the green line and the top of the red bar). Table 2 provides details on infant-only clusters.

**Table 2.** Infant only clusters (age < 1 year)

|  |  |
| --- | --- |
| Total number of infant cases | 21019 |
| Total number of infant cases in clusters ≥ size 2 | 7994 |
| Total number of infant-only clusters ≥ size 2 | 1366<br>(16.7%) |
| *Largest size infant-only clusters* | *1 cluster of size 8*<br>*(all clusters $\geq 4$ are single state)* |

Infant only clusters (age < 1 year)

**Table 3.** PulseNet clinical isolates in study set.

|  |  |  |
| --- | --- | --- |
| *Salmonella* | 1961-08-23 : 2023-10-27 | 265,449 isolates<br>>98% from<br>2015-01-01 |
| *Escherichia coli* | 1981-01-02 : 2023-11-04 | 68,527 isolates<br>>96% from<br>2015-01-01 |
| *Campylobacter* | 1983-01-01 : 2023-11-07 | 23,577 isolates<br>>95% from<br>2015-01-01 |
| *Listeria* | 1965-11-20 : 2023-12-152 | 8,205 isolates<br>>90% from<br>2013-01-01 |

Because we want to focus on the possibility of infant formula as a source of contamination, we are including those individuals less than 1 year old rather than < 6 months old because this is the age in which most children transition from infant formula to cow’s milk. There is only one cluster at the maximum size of 8 cases and all clusters >= size 4 are single state clusters. Both of these points provide strong support for the conclusion that *Salmonella* contamination of infant formula *at the production site* could only be responsible for a very small fraction of cases, if any, and any possible contamination episodes from this source would be exceedingly small.

If infant formula contaminated at the site of production is not a major source of salmonellosis then what *is* the source of contamination for cases at age < 6 months? The diets of infants under 6 months of age are largely restricted to infant formula and breast milk, neither of which is commonly consumed by individuals of age 10 or older. Co-occurence of these age groups in a cluster therefore suggests cross-contamination between non-infant food and infant formula or breast milk or exposure to a common non-food source of contamination. Figure 13 shows the *Salmonella* case counts for various age ranges of children and the number of cases in that age range which are in clusters along with cases with age at least ten years. Unsurprisingly, since Figure 12 shows that only a small fraction of the infant (age ≤ 6 months) cases are in infant-only clusters, Figure 13 shows that most of these infant cases cluster with cases from individuals ≥ 10 years of age. The pattern for the other age ranges, where their dietary intake is more similar to older children and adults, is also quite similar.

**Figure 13.**
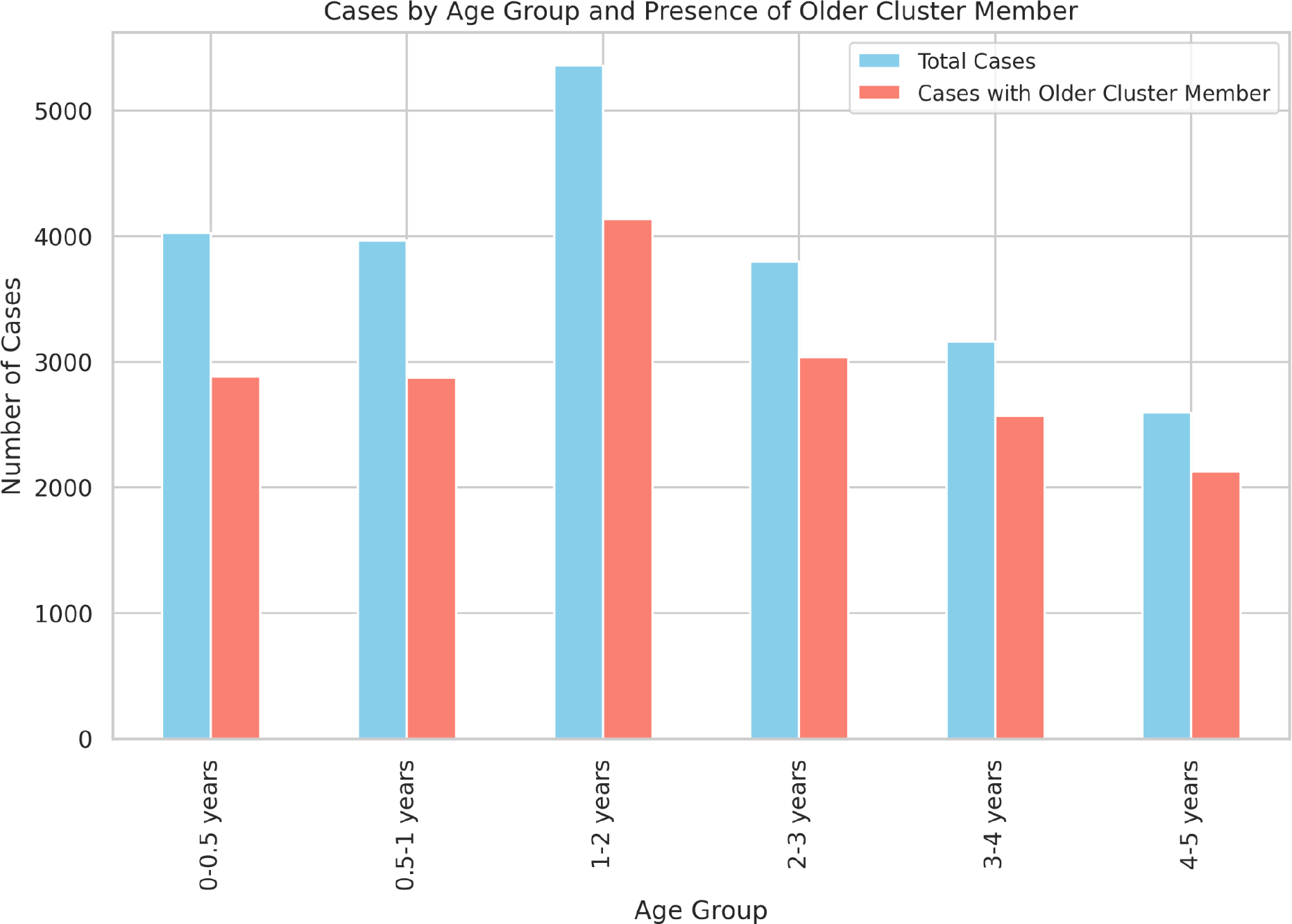
S*a*lmonella case counts by age range and when clustered with case ≥ 10 years old *Salmonella* case counts in non-singleton clusters by age range and when clustered with cases aged ≥ 10 years.

Patterns similar to those in Figure 12 and 13 and Table 2 are evident in *Campylobacter* and *Escherichia coli* (data not shown).

## Methods

### Data sets

The pathogen isolates used in this project were collected and sequenced by CDC’s PulseNet national laboratory network for foodborne outbreak detection. The pathogen genome data and SNP distances were downloaded from the National Center for Biotechnology Information (NCBI) Pathogen Detection site (“Home - Pathogen Detection - NCBI” n.d.). The identifiers for the pathogen isolates used for these analyses are available in spreadsheets listed in the Supplementary files (PDT*) along with identifiers for the clusters and the size of each cluster based on thresholds of 2, 4, and 8 SNPs. For *Salmonella*, the serovars as identified in the NCBI pipeline are listed as well. The Pathogen Detection releases used for each pathogen are:

● *Campylobacter* PDG000000003.2084 (11/2023)
● Ecoli_Shigella PDG000000004.4162 (11/2023)
● *Listeria* PDG000000001.3486 (11/2023)
● *Salmonella* PDG000000002.2848 (11/2023)

### Metadata

The PulseNet metadata used for these analyses was accessed from CDC’s SEDRIC database under their data-use agreement (“SEDRIC: System for Enteric Disease Response, Investigation, and Coordination” 2022). The fields used were:

● the age of the affected individual,
● the date of collection, and
● the state where the isolate was collected.

### Generating clusters

#### SNP distances

The SNP distances for these analyses were the patristic (i.e. tree-based) distances computed by the NCBI Pathogen Detection resource (“Home - Pathogen Detection - NCBI” n.d.).

This resource has been available since 2016 and has been used intensively by the FDA, CDC, USDA, and US state public health laboratories for outbreak detection and source tracking. A summary of the pipeline is available (https://ftp.ncbi.nlm.nih.gov/pathogen/Methods.txt).

#### SNP clustering

There are many ways to generate clusters using distance measures such as SNP distances. We start with the NCBI Pathogen Detection SNP clusters that are updated regularly. These SNP clusters include all genomes that are within 50 SNPs of each other (see NCBI pipeline summary https://ftp.ncbi.nlm.nih.gov/pathogen/Methods.txt). We determine the case cluster for the genomes of each PulseNet clinical isolate within an NCBI SNP cluster as follows: Using single linkage clustering based on a threshold SNP distance, isolates are added to a case cluster as long as they are less than or equal to the threshold SNP distance from any of the isolates in the cluster. Any isolate which is further away than the threshold from all other isolates would be in a cluster of size 1.

#### SNP thresholds

Choosing a SNP threshold that implies that two isolates share a similar source of contamination in this analysis has been discussed in the context of outbreak detection and source tracking (Pightling et al. 2018; Chattaway et al. 2023) In this analysis, we are using the SNP threshold to generate clusters from very large sets of clinical isolates and then examining statistical trends in the metadata associated with these clusters. To understand the robustness of the results, we compared thresholds set at 2, 4, and 8 SNPs for most analyses. Figure 14 shows the cumulative frequency curves for *Salmonella* (similar to Figure 1 though this is for all serovars including Enteritidis) and as one would expect, a threshold of 2 shifts the curve towards smaller clusters (i.e. a higher frequency at cluster size 1) and a threshold of 8 shifts the curve towards larger clusters (i.e. a lower frequency at cluster size 1). For all three thresholds however, the majority of cases are in smaller clusters (e.g. < size 10).

**Figure 14.**
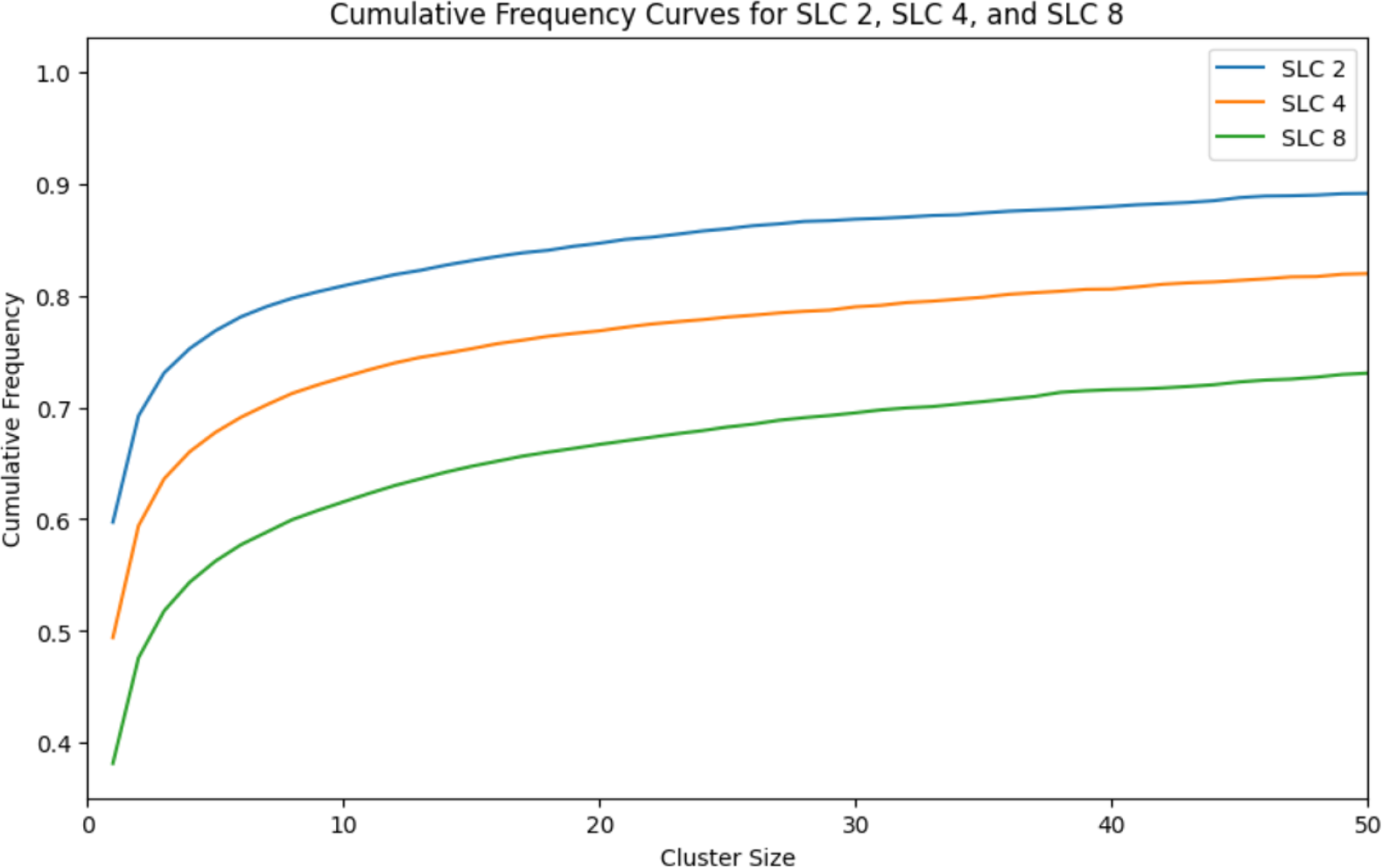
Cumulative frequency curves for *Salmonella* using SNP thresholds of 2, 4, and 8 SNPs Cumulative frequency curves for *Salmonella* using SNP thresholds of 2 (blue), 4 (orange), and 8 SNPs (green).

The impact of these different thresholds on the observations in Figure 2 showing the bias of younger age groups towards smaller clusters is shown for *Salmonella* in Figure 15 and again, this would not change the interpretation.

**Figure 15.**
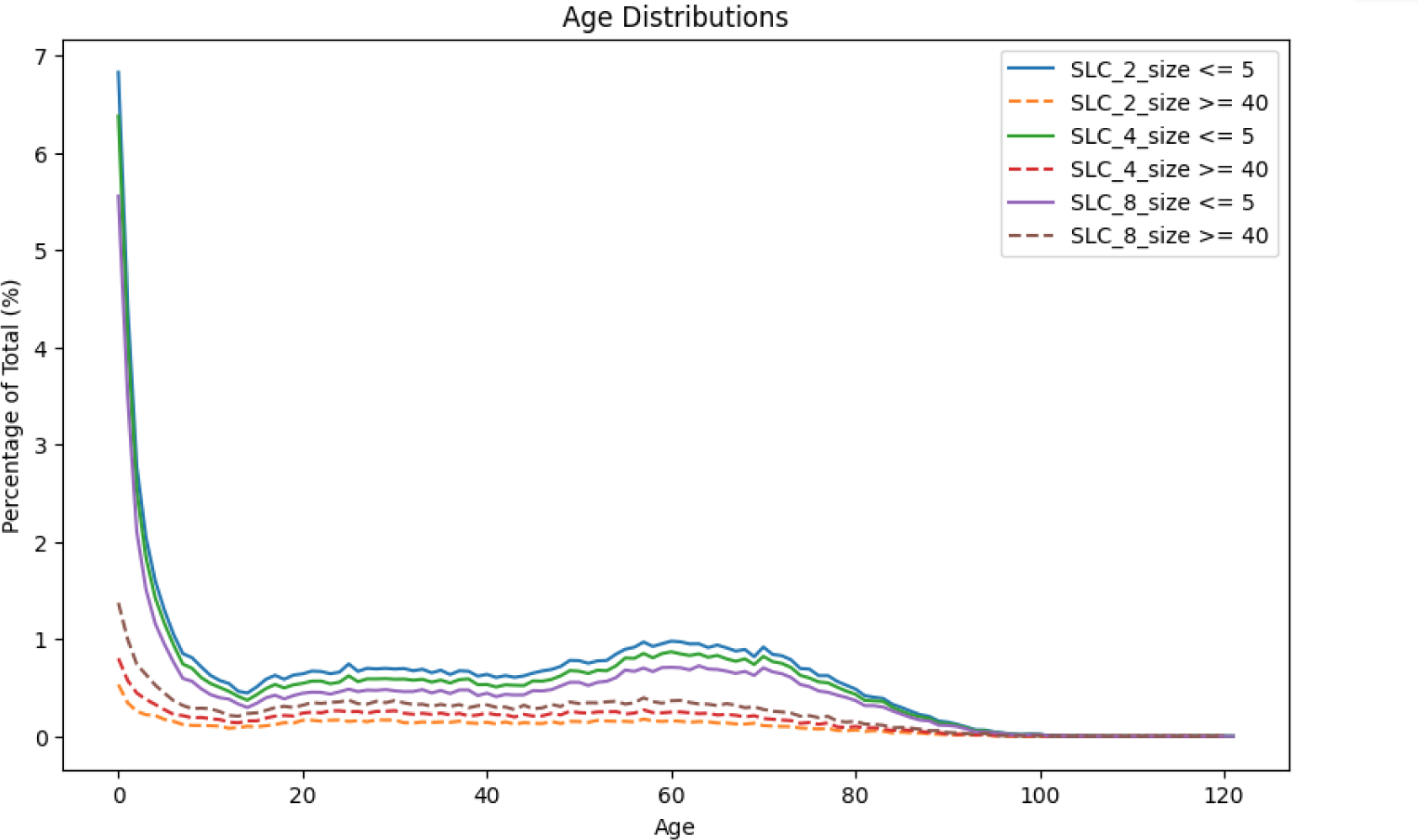
Age (unadjusted) distributions for small and large cluster ranges using SNP thresholds of 2, 4, and 8 SNPs Comparison of the percent of cases in different age groups within small and large cluster sizes and using SNP thresholds of 2, 4, and 8 SNPs to form the single linkage clusters. These are actual case counts by age and are not normalized for the demographics.

The shifting of the cluster size distributions by the SNP thresholds affect some analyses in obvious ways. As one increases the threshold, the fraction of multi-state *Salmonella* clusters (Table 1) increases from 74% (2 SNPs), to 78% (4 SNPs), to 83% (8 SNPs). The median persistence for *Salmonella* clusters (Figure 10) increases from 469 days (2 SNPs), to 1071 days (4 SNPs), to 1982 days (8 SNPs). Either way, the interpretation is the same - i.e. that even though most cases are due to small contamination episodes, a high fraction of these contamination episodes are multi-state and highly persistent. It is perhaps even more surprising that half of the clusters as tight as those generated using a threshold of 2 SNPs persist for over 469 days. It was noted above that Enteritidis clusters formed using low SNP thresholds may still lump together multiple independent proximal contamination episodes (Baker et al. 2023; Dallman et al. 2016). This merging of independent proximal contamination episodes by Enteritidis could impact the persistence analysis for *Salmonella*. When Enteritidis is excluded from the dataset, the median persistence duration for *Salmonella* clusters decreases from 1071 to 705 days, indicating a substantial reduction yet affirming the high persistence characteristic of these clusters.

For analyses of the composition of clusters such as those in Figure 13, the SNP thresholds have a smaller impact but may not be monotonic - e.g. the fraction of infants ≤ 6 months old in clusters with individuals ≥10 years increases slightly from 2 SNPs to 4 SNPs and then decrease slightly at 8 SNPs.

## Discussion

Most of what we know about the contamination episodes that cause the estimated 48 million cases of food poisoning each year in the United States are from the small fraction of cases in the foodborne outbreaks that are investigated by the CDC, FDA, USDA-FSIS, as well as state and local health authorities (Scallan et al. 2011). The vast majority of food poisoning cases are classified as sporadic cases which have primarily been studied through case-control methods as well as routine surveillance systems (Devleesschauwer et al. 2019; Fullerton et al. 2012; Domingues et al. 2012; Ebel et al. 2016). By analyzing the clusters formed from closely related pathogen genomes of clinical isolates within CDC’s PulseNet system, we can derive a more detailed picture of the contamination episodes underlying foodborne illness in the United States.

The clusters of a given size shown in Figure 1 are a mixture of contamination episodes with a range of different sizes since, as noted in the Introduction, only a variable fraction of the cases would be reported within the PulseNet system. In addition, the SNP threshold used to generate the clusters may be too stringent in some instances and thus split cases from the same contamination episode into different clusters, while in other instances it may be too high and thus merge cases from different episodes. Note also that some US states began the use of whole genome sequencing before others so the coverage is more comprehensive later in the time interval of the sample set (see Methods). Furthermore, since we are likely to be missing cases from contamination episodes towards the beginning and ends of the time interval corresponding to our sample collection, the clusters associated with these episodes will be incomplete.

Despite these caveats, the results presented here demonstrate substantial differences in the epidemiological properties of the clusters of different sizes, i.e., there are strong signals associated with the computed clusters.

The assumption that clinical isolates separated by a small number of SNPs generally share the same proximal source of contamination may be less likely for Enteritidis than the other foodborne pathogens. Eggs and poultry are the most common source of Enteritidis cases in the United States (Jackson et al. 2013). Because the same breeder site may supply multiple poultry production facilities, identical or nearly identical strains may be found at these separate production facilities (Zhang et al. 2021; La et al. 2021; Lei et al. 2020). Thus, Enteritidis clusters formed using low SNP thresholds may lump together multiple independent proximal contamination episodes and the interpretation of the Enteritidis results must account for that possibility (Baker et al. 2023; Dallman et al. 2016).

We see a consistent pattern for *Campylobacter*, *Escherichia coli*, and *Salmonella* :

● The smallest contamination episodes account for the largest fraction of cases, except for Enteritidis (Figure 1);
● The diversity decreases, e.g., in terms of serovars for *Salmonella*, with increasing cluster size (Figure 4);
● the case rates are higher in the younger age ranges in both large and small clusters, albeit with the drop off noted at ages < 1 year (Figure 2);
● The youngest age groups, and to a lesser extent the oldest, are most overrepresented in the smallest clusters (Figures 2 and 5).

The last finding is consistent with earlier work (Ebel et al. 2016) that found that the youngest age groups were underrepresented in *Salmonella* outbreaks as compared with sporadic cases (i.e., in general, smaller clusters).

One possible explanation for the dramatically higher case rate among the youngest children is the greater likelihood of seeking medical care and thus being picked up in the PulseNet system. It is not clear, however, how this factor would explain the differences seen in Figure 2 between smaller and larger contamination episodes.

Another possible explanation would be based on age-based differences, in terms of exposure or susceptibility, to different genetic subgroups within the pathogen species, e.g. different *Salmonella* serovars. Consistent with this possibility, the composition of the clusters changes with cluster size - with increasing diversity of serovars or genetic subgroups in the smaller clusters (Figure 4). However, we see the increased case rates for the youngest age groups (and to a lesser extent, the oldest age groups) among both the common and less common *Salmonella* serovars (Figure 5). And even Enteritidis, which is unique in its bias for larger clusters, follows the same pattern of relatively higher rates in the smallest clusters for the youngest and oldest age groups.

A simple explanation that would integrate many of the reported findings for *Salmonella*, *Escherichia* coli, and *Campylobacter* is based on two assumptions:

1. Susceptibility is age-related, with the youngest age groups, and to a lesser extent, the oldest having greater susceptibility;
2. There is a distribution of contamination levels in, e.g., food servings, associated with a contamination episode: episodes that contaminate smaller numbers of servings will, on average, generate a smaller fraction of servings with pathogen levels sufficiently high to be likely to cause symptoms in most individuals.

Regarding Assumption #1, others have noted that the youngest age groups were more susceptible to foodborne illness (Thomas et al. 2015). This is perhaps expected given our knowledge about the maturation of the immune system cf (Simon, Hollander, and McMichael 2015; Georgountzou and Papadopoulos 2017)). Note that the drop in infant cases at the very youngest age group may relate to some level of protection from maternal antibodies (de Alwis et al. 2019; Basha, Surendran, and Pichichero 2014). Another reason for the drop may relate to changes in the fraction of infants that are exclusively breast-fed: at birth the rate is 63%, by 3 months the rate is 45%, and by 6 months, the rate is 25% (CDC 2023).

It seems reasonable therefore to assume that there would be a distribution of pathogen levels in servings associated with a contamination episode (assumption #2). While a large contamination source might produce a substantial number of servings with a sufficiently high level of pathogen to sicken healthy individuals, such a source would likely produce a number of *additional* servings with dosages that are only likely to cause symptoms in more susceptible consumers. Likewise, a smaller contamination source may yield a lower fraction of servings with pathogen levels sufficiently high to reliably cause symptomatic illness in most individuals. Because of the increased susceptibility of the younger age groups, a higher fraction of the contaminated servings are able to produce symptomatic illness in this age group, producing the age incidence curve in Figure 2. Furthermore, this difference would be more pronounced for the smaller clusters if the distribution of pathogen levels per serving were shifted according to the second assumption.

Note that this explanation for *Salmonella*, *Escherichia coli*, and *Campylobacter* is mostly based on the phenotype of the host, i.e., that *overall*, susceptibility is primarily age-related, along with a very general property of contamination episodes (i.e. assumption #2) rather than on the phenotype of the pathogen or other factors. Listeria however looks quite different and thus the phenotype of the pathogen can lead to very different age incidence.

Of the major *Salmonella* serovars, Enteritidis also stands out with a lower bias towards younger age groups and less difference in this bias between larger and smaller clusters. The latter may relate to the greater likelihood, as compared with other serovars, of merging separate contamination episodes of Enteritidis noted above. However, the possible increased merging of clusters would not explain the overall decrease in age differences. Perhaps the susceptibility of younger age groups is only slightly increased for Enteritidis or some aspect of its phenotype weakens the relationship between the number of contaminated servings and the concentration of pathogen. In conjunction with the observation that a high fraction of infant cases of *Salmonella* are due to cross-contamination in the home, reduced environmental persistence could lead to incidence in younger age groups that is closer to adult incidence. Possibly relevant here is the observation that seasonal variability of Enteritidis is also markedly reduced compared to the other major serovars (see e.g. (“*Salmonella* Atlas” 2020)). Since poultry is the main source for Enteritidis we compared the incidence by age for clinical isolates of Enteritidis within 2 SNPs of poultry isolates with that of clinical isolates of other serovars also linked to poultry. The Enteritidis distribution was similar to that seen on Figure 3 while clinical isolates of other serovars similarly linked to poultry showed the same increased age bias seen in Figure 2a (data not shown).

This relationship between the age distribution of cases and cluster sizes can be helpful in exploring geographical differences among *Salmonella* serovars as seen in the CDC surveillance data. For example, the FoodNet data for available states (i.e., Georgia as compared with New York and Connecticut) shows that the fraction of *Salmonella* cases among the youngest age groups may be higher in the Southeastern US as can be clearly seen in Figure 5a (“FoodNet” 2023). The national surveillance data show that levels of Enteritidis relative to other serovars are higher in the Northeastern and lower in the Southeastern US and Hawaii (as seen in Figure 7a) and the opposite is true for the Newport serovar (“2016-Salmonella-Report-508.pdf,” n.d.).

These observations can perhaps be better understood in the context of the observation (Figure 7c) that the fraction of cases in cluster size 1 is higher in the Southeast and Hawaii. In other words, smaller contamination episodes with lower average contamination levels are more common in the Southeast and Hawaii and thus there is a higher fraction of infant cases (perhaps because of greater susceptibility) and a lower fraction of Enteritidis cases. While it seems plausible that these differences in the Southeast and Hawaii are climate-related, i.e., these states have warmer, more humid weather, the fact that infant fraction is not seasonal, even within a state, suggests that the phenomenon is not a simple consequence of climate (Figure 8 and data not shown). However, the Southeast and Hawaii also have a higher fraction of cases in single state clusters (Figure 7d) which would be consistent with local environmental exposure, perhaps from reptiles that thrive in these regions, c.f. (Pees et al. 2023).

Although the smallest contamination episodes account for the majority of clinical cases of food poisoning, these do not appear to be primarily local, like some sort of environmental exposure or associated with exposure from a restaurant outbreak. Rather, a majority of these cases are in geographically dispersed clusters (Table 1). For example, 78% of *Salmonella* cases (cluster size >=2) are in multi-state clusters.

This implies that the servings that cause these cases are being distributed from central sites where the contamination is occurring, which would primarily be commercially distributed foodborne transmission, but could also include contact with commercially distributed animals/pet food, and returning travelers. Furthermore, a high fraction of cases are in clusters that are persistent, e.g., half the *Salmonella* cases are in clusters that persist for over 1071 days. As discussed in the Methods section, the results on the fraction of multi-state clusters and on persistence of clusters is dependent on the choice of SNP thresholds for the generation of the clusters however the qualitative picture remains the same: over a wide range of thresholds, the majority of *Salmonella* cases are the result of contamination distributed from central sites and persisting for extended periods of time. As noted above, because we are missing isolates that could be potential members of clusters prior to the beginning and after the end of our sample window as well as the larger number of cases that are not detected e.g. by PulseNet, these are likely to be conservative estimates.

If infant formula contaminated at the production facility were a significant cause of salmonellosis, we would expect to see a substantial fraction of infant-only clusters (size ≥ 2). However, they represent only 17.6% of the cases. Furthermore, almost all infant-only clusters are single-state: there are only 2 infant-only clusters at the maximum size of 8 cases, and both are single-state clusters (Figure 12 and Table 2). Rather, as seen in Figure 13, it appears that most cases of salmonellosis in infants (age < 6 months) are due to cross-contamination at home because they co-occur in clusters with individuals too old to consume infant formula. While the published literature on the importance of cross-contamination versus undercooking does not have a consistent message (Luber 2009; Møretrø et al. 2021), our results suggest that at least for the youngest age ranges, cross-contamination within the home is the major cause of salmonellosis.

While our results imply that most cases of foodborne illness are due to contamination distributed from central sites over an extended period of time, these cases are due to many very small contamination episodes and the levels of pathogen at the central sites would likely be extremely low. These very low levels of contamination may be difficult to detect, e.g., in a production facility or the environment, and even more difficult to eliminate. This may help explain the slow progress in reducing the overall burden of foodborne illness in the United States (“FoodNet” 2023). Given the high case rates of the youngest age groups, their increased susceptibility, and the likelihood that a high fraction of these cases are due to cross-contamination, a greater emphasis on improving food safety in the consumer household should be considered. While improving food safety practices in the household would be quite challenging (“Kitchen Life 2” n.d.) identifying the minimal changes needed to reduce cross-contamination of infant formula may be more feasible. Currently state laws require infant car seats and hospitals provide training on installing and using a car seat for parents bringing their newborn home from the hospital. While breastfeeding is encouraged, much less attention is given to educating new parents and caregivers on the safe preparation of infant formula and the use of feeding bottles (Redmond and Griffith 2009).

Infection from environmental sources rather than food may also contribute to higher case rates in the young. Among *Salmonella* types with especially high overrepresentation in infants, environment-associated types, including types found mainly in reptiles, predominate (Figure 6 and Supplementary Figure 2). Some geography-related observations might also be explained by this phenomenon: the higher fraction of same-state cluster members among infants (due to the local source of infection) and the higher fraction of infant cases in the southeast (due to greater reptile abundance).

*Salmonella* Enteritidis seems to be a more feasible target for improving food safety outside the household than the other major pathogens because the contamination episodes causing most cases are larger and a significantly higher fraction of these are likely to have been distributed widely from central sites.

Moreover, we know the primary source of *Salmonella* Enteritidis: poultry and eggs (Gould et al. 2013). Quantitative risk assessment models have been created based on the pathogen survival rates in the cooking process that have fairly good correlations between the prevalence of *Salmonella* on, e.g., poultry and the fraction of cases in the US (Oscar 2004; Rajan, Shi, and Ricke 2017). And while Enteritidis may, in principle, be a more feasible target for risk reduction at central sites, improved food safety practices in the household would be helpful here as well.

While WGS has already proven to be a useful tool for identifying and investigating foodborne outbreaks, we have demonstrated that the increasingly comprehensive set of pathogen genomes can also reveal important aspects of sporadic foodborne illness which accounts for most cases of food poisoning.

## Supporting information

Supplementary Figure 2 (HTML)

Supplementary Cluster files

## Data Availability

The PulseNet metadata used for these analyses (state, collection date, and age) was accessed from the CDC SEDRIC database under their data-use agreement. All other data is contained in the manuscript and supplementary data.

## Acknowledgments

The authors wish to thank the PulseNet participating laboratories for isolating and sequencing the *Salmonella, Listeria, Escherichia coli, and Campylobacter* isolates, uploading the sequences to the PulseNet National Database, and submitting the raw sequence data to the NCBI public databases. We also thank the CDC epidemiologists for linking isolates together during outbreak investigations and their feedback on the manuscript. This work was supported in part by the intramural research program of the National Library of Medicine, National Institutes of Health. The opinions expressed in this article are those of the authors and do not reflect the view of the National Institutes of Health, the Department of Health and Human Services, or the United States government.

