## Supplementary material for "Genomic perspectives on foodborne illness": Supp Figure 1

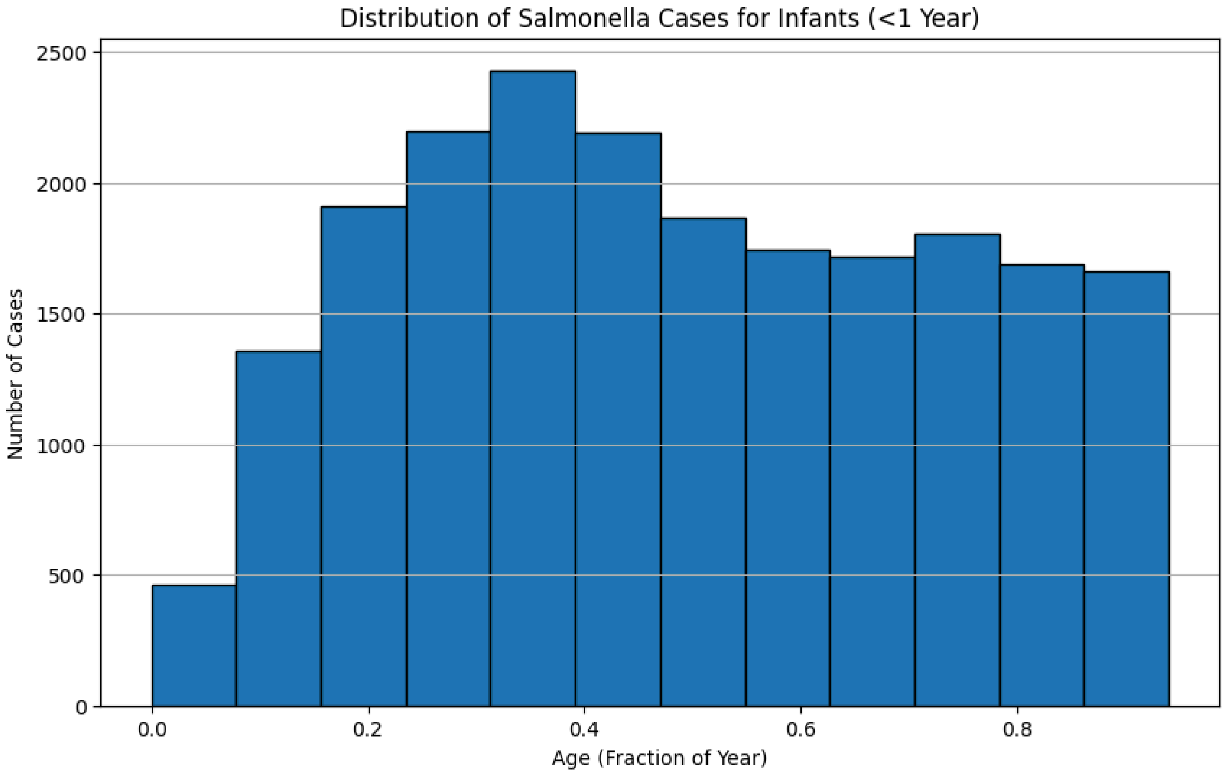


Supplementary Figure 1

Number of Salmonella cases for infants < 1 year of age in fractions of one year.
